# The Ultra Fit Community Mask - Toward Maximal Respiratory Protection via Personalized Face Fit

**DOI:** 10.1101/2021.07.01.21259428

**Authors:** Chulho Hyun, Mark Martin Jensen, Kisuk Yang, James C. Weaver, Xiaohong Wang, Yoshimasa Kudo, Steven J. Gordon, Jeffrey M. Karp, Anthony E. Samir

## Abstract

Effective masking policies to prevent the spread of airborne infections depend on public access to masks with high filtration efficacy. However, poor face-fit is almost universally present in pleated multilayer disposable face masks, severely limiting both individual and community respiratory protection. We developed a set of simple mask modifications to mass-manufactured disposable masks, the most common type of mask used by the public, that dramatically improves both their personalized fit and performance in a low-cost and scalable manner. These modifications comprise a user-moldable full mask periphery wire, integrated earloop tension adjusters, and an inner flange to trap respiratory droplets. We demonstrate that these simple design changes improves quantitative fit factor by 320%, triples the level of protection against aerosolized droplets, and approaches the model efficacy of N95 respirators in preventing the community spread of COVID-19, for an estimated additional cost of less than 5 cents per mask with automated production.

**Teaser:** A personalizable, low-cost mask improves facial fit, reduces user exposure, and decreases spread of contagious aerosols.

## Introduction

The COVID-19 pandemic caused by the severe acute respiratory syndrome coronavirus (SARS-CoV-2) remains an unprecedented threat to global public health. SARS-CoV-2 is known to spread via respiratory droplets and aerosols asymptomatically and pre-symptomatically (1). In the absence of cure or vaccination, numerous non-pharmaceutical public health interventions such as travel restrictions, mass screening, quarantine centers, and contact tracing have been implemented to control the pandemic (2). Even after successful vaccination, masks can be protective while resistance develops, prevent potential asymptomatic spread, and provide protection for immunocompromised individuals and in communities where vaccination has been limited. (3)

As a low-risk, precautionary control method (4), universal face masking has shown effectiveness in reducing disease spread (5). However, the performance of most cloth and disposable 3-ply, 3-pleat disposable masks (3PM) is degraded by poor face fit, resulting in leakage of aerosols at gaps between the mask and a wearer’s face. This leakage reduces the quantifiable ‘fit factor’, a metric of mask fit performance. Conventional 3PM have fit factors (FFs) of only 2.6 - 4.4 (6, 7), equivalent to ca. 20 - 40% leakage. A further limitation of conventional disposable masks occurs during coughing and sneezing, which elevate the mask, enlarging peripheral gaps. To this effect, disposable masks work least well when source control is the most required.

Respirators, such as the surgical N95 respirators, fit tightly and provide sufficient protection from fluid and airborne pathogens but were designed for short-term use in high risk environments, not in community settings. The N95 respirators are of shortage, require fit testing, and are uncomfortable to wear for extended periods, which leads to poor adherence, frequent mask removal, and even personal injury (8,9,10). Barrier face coverings offer less protection but are better tolerated by users due to comfort and ease of use (11). Due to large variability in the performance of commercially available consumer masks, the American Society for Testing Materials (ASTM) and National Institute for Occupational Safety and Health (NIOSH) released new standards from masks designed provide community and workplace protection against the spread of airborne contagion (12,13). A central feature of these new standards is their requirements for fit, breathability, and filtration.

Poor mask fit is due to 1) mismatch between mask and face shape, and 2) looseness of the mask, usually due to inability to adjust mask tightness. A variety of secondary devices (e.g. mask fitters, extender straps) and techniques (e.g. knotting, tucking, double masking) have been developed to address these limitations, but have not been broadly adopted, and many exhibit effectiveness by compromising comfort and convenience (14, 15).

A simple and efficacious face covering technology would therefore likely promote broader adoption and improve user experience and compliance.

To overcome the the lack of protection offered by current face masks and avoid the discomfort generated from respirators, we developed a novel *Ultra Fit Mask* (UFM) by adding three innovations to the conventional 3PM design: (1) a peripheral moldable element, (2) an integrated ear loop tension adjustment, and (3) an inner flange element designed to trap exhaled aerosols. Each design element was specifically considered for its ability to be incorporated into current automated mass manufacturing systems with minimal modification and cost. By incorporating easy-to-manufacture features directly into the designs of mass-produced disposable masks, we enable rapid implementation and maximize the potential impact of these design innovations.

## Results

### Ultra Fit Mask - Improved Mask Design

To facilitate improved fit and seal gaps, we added a moldable wire element along the entire periphery (Fig. 1A, 1B) of a conventional 3-ply disposable mask (3PM). With the moldable element, the mask gains a pliable structure that can conform to any face, with minimal gaps. Compared to the conventionally used 100 mm-long nose bridge wire, a longer and thicker nose wire (length > 120mm, diameter > 0.58mm) provides a better seal across the entire nasal-infraorbital region (bizygomatic breadth). The wires on the cheek and the chin edges of the mask provide a fitting experience similar to cup-shaped respirators and enable customized shaping of the mask edge. Pinching the side or chin wires reduces the effective length of the sides which tightens the fit and further reduces gaps (Fig. S1), and as such, these face-conformal adjustments improve fit, particularly for users with smaller faces.

**Figure 1.**
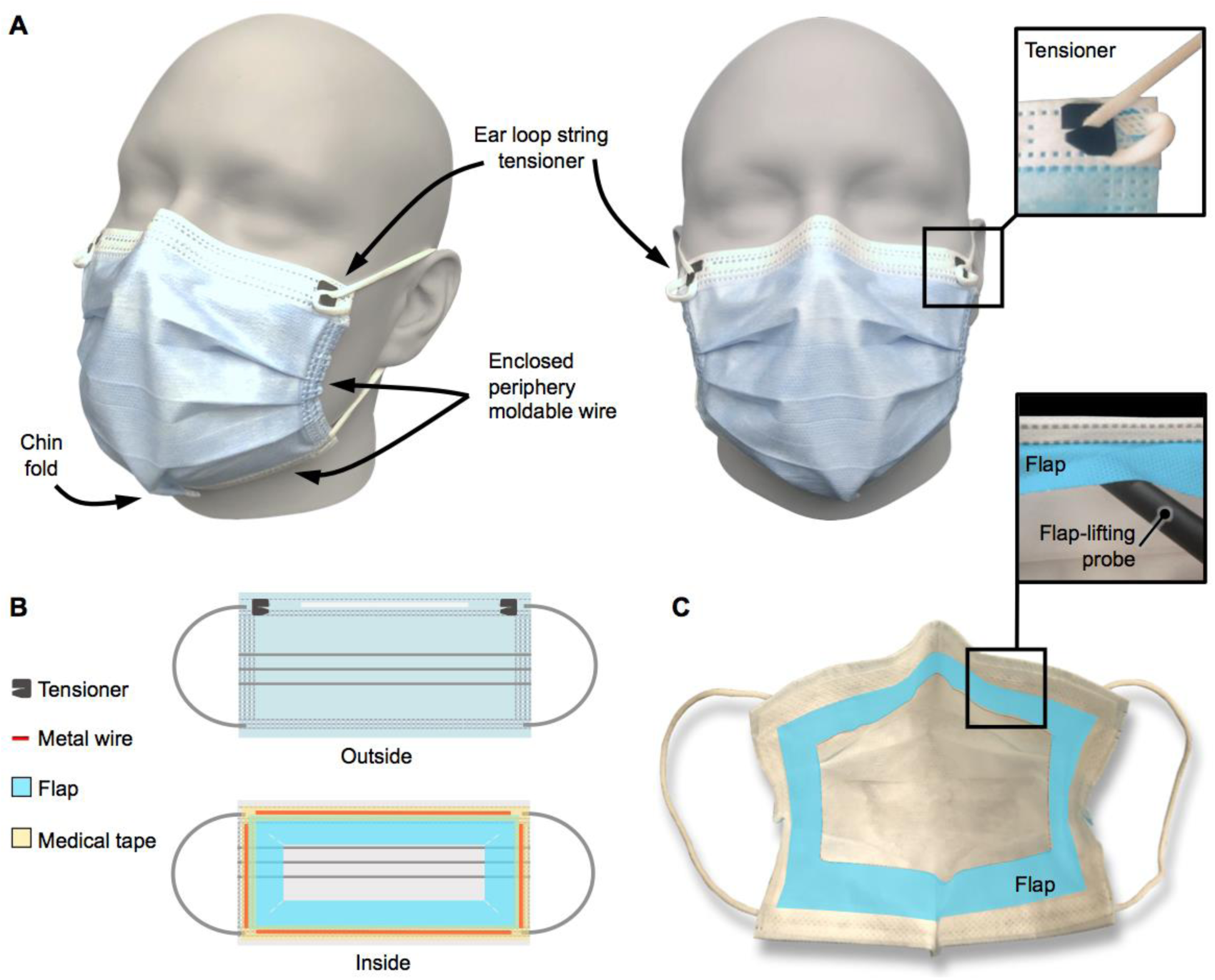
Ultra Fit Mask for Reducing Face Seal Leakage on the Periphery of the Mask Body. (A) Ultra Fit Mask (UFM) fitted on a head form compliant to ISO 16976-2 Respiratory Protective Devices (medium size). (B) Exterior and interior schematics of prototype UFM. (C) Additional cough-trapping internal flap layer attached on the interior side is highlighted in blue.

To further promote a tight face seal, we added an adjustable tension element to the ear loops. In conventional 3PM, elastic ear loop strings compress the mask onto the wearer’s face, with mask-to-face compression largely a function of wearer head size rather than comfort or face seal. Integrated UFM tension adjustment components cinch the elastic ear loop strings and create user-adjustable mask-to-face compression. Appropriate tensioning in combination with the face-conforming mask shape maximizes both comfort and face seal (Fig. 1A).

To prevent the escape of droplets via the sides of the mask, we added an inner flange as a “droplet trap” (Fig. 1C). Coughing generates pressure that displaces the mask away from the face, reducing face seal exactly when it is needed most to capture rapidly ejected respiratory droplets (the flange serves to capture these droplets).

### Qualitative Assessment

#### Exhaled Vapor

To evaluate the ability of the masks to reduce aerosol spread, we analyzed video recordings of vapor exhalation via laser light scattering as a qualitative assessment of mask fit. To image escaping vapor, video recordings were taken from the side and the front of a wearer (Fig. 2A,C). Vapor escaping through open gaps for the 3PM was visible near the forehead, while vapor release through the side of the mask was clearly visible as a gray plume near the ears (Fig. 2A, red arrow). Qualitative assessment of UFM vapor unequivocally demonstrated less vapor escape, with more vapor instead being forced through the mask filter material (Fig. 2A, yellow arrow). These observations were confirmed by plotting relative laser scattering (RLS) of every frame of the acquired videos over time (Fig. 2B,D). Leaked vapor from the UFM at the nasal-infraorbital side was nearly an order of magnitude less for the first 1.5 seconds than that with the 3PM (Fig. 2B). After 1.5 seconds, a higher RLS with the UFM was observed, corresponding to scattering of floating vapor that penetrated the mask filter material earlier (Movie S2). The UFM demonstrated negligible cheek leakage, with only a small leak plume arising from the chin region (Fig. 2C, yellow arrow). With the UFM, the amount of laser scattering from around the cheeks due to leakage was reduced by ca. two orders of magnitude.

**Figure 2.**
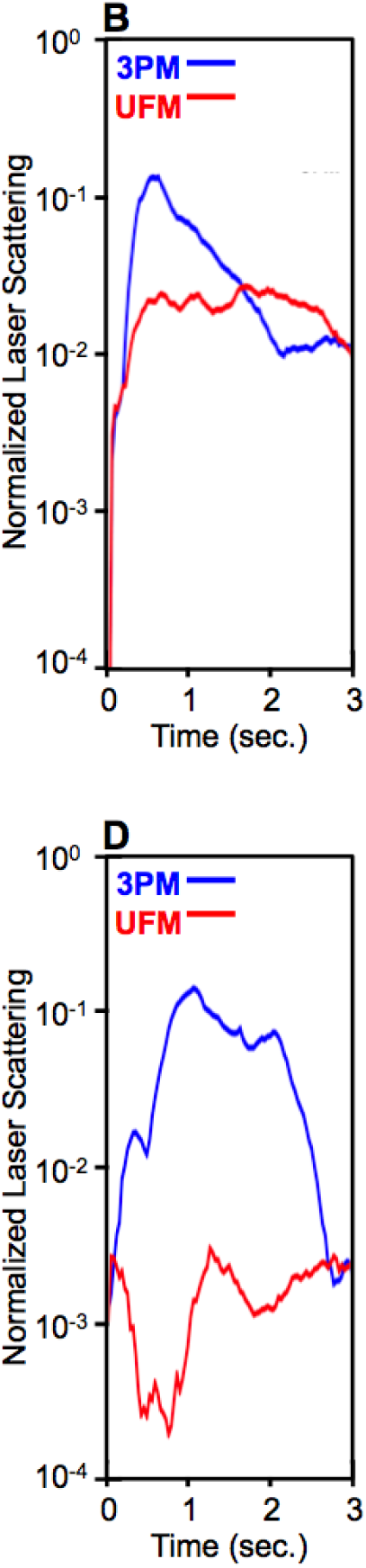
Evaluation of Exhaled Vapor using Laser Scattering. (A,C) Side-view and front-view exhaled vapor imaging and histograms of the segmented area (white dotted box). Red arrows indicate particles that escaped the mask, while yellow arrows indicate particles that passed through the mask filter material. (B,D) Normalized laser scattering of the segmented forehead and lateral regions over an exhalation period. (C) Front-view exhaled vapor imaging and histogram of the segmented area (white dotted box). ***NOTE:*** *Per medRxiv policy that manuscripts not contain ‘photographs/videos and any other identifying information of people’ we have removed panels A&C and movie S2 to comply with this policy. In the these items, an author of this manuscript can be seen wearing the mask as part of testing. The full figure is available upon request to the coresponding author or via the following link*. https://www.dropbox.com/sh/nkg2k1gu4aih3wz/AADy_o3CqlovLpCuXTE44uCYa?dl=0

#### Infrared Thermal Imaging

To further assess leakage sites for both inhalation and exhalation, we used infrared thermal imaging during normal breathing in an air-conditioned room. Synchronous thermal changes at the mask surface with breathing cycles were visible as relatively cool inhaled air and relatively warm exhaled breath pass through the mask filter material. Inward and outward leakage along the nose-infraorbital edge of the mask were visualized as zones of high thermal variation over time (Fig. 3A). The ingress of cool air through the nasal-infraorbital edge of the mask at the end of inhalation and egress of warm breath at the end of exhalation indicated leakage for the 3PM. For the UFM, the nasal-infraorbital edge remained relatively warm during breathing cycles except for a small ingress on the right side of the wearer (Fig. 3A). Over a 2-minute breathing cycle, the UFM showed larger temperature variation in the center of the mask consistent with cycling of cool inhaled air and warm exhaled air traversing the mask filter material (Fig. 3B) Thermal variation analysis at the right and left sides of the nasal-infraorbital edge of both masks showed that a sinusoidal response in the UFM was less pronounced. A higher offset temperature around the edge of the UFM was due to increased skin contact owing to the reduction in mask edge gaps (Fig. 3C). Overall, thermal imaging demonstrated that the UFM directed air through the filter material and removed ingress and egress air leaks around the mask edge.

**Figure 3.**
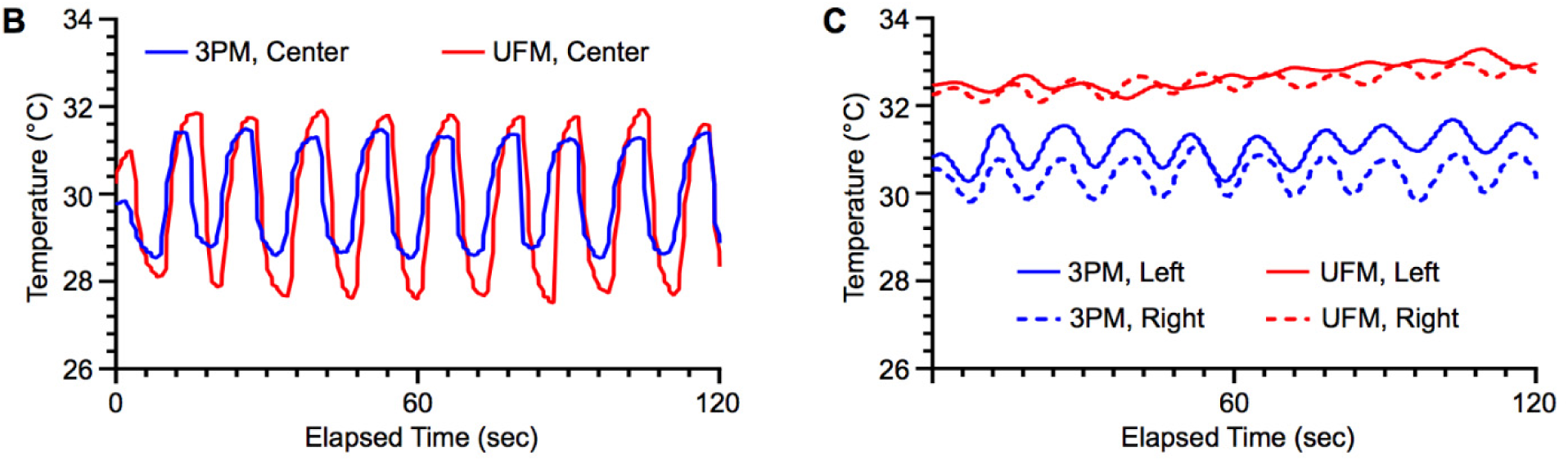
Infrared Thermal Imaging of Mask Surface over Breathing Cycles. (A) Infrared thermal snapshots at the end of inhalation and exhalation with 3PM and UFM were obtained from video recordings of breathing cycles. Three subregions at the left and right side of the nose (red dotted boxes, 5 × 5 pixels), and the center of the mask (red dotted box, 20 × 20 pixels) were selected for thermal variation analysis over multiple respiration cycles. Thermal variation of the (B) center subregion and the (C) left and right side of the nose over a 120-second breathing cycle. ***NOTE:*** *Per medRxiv policy that manuscripts not contain ‘photographs/videos and any other identifying information of people’ we have removed panel A and movie S3 to comply with this policy. In the these items, an author of this manuscript can be seen wearing the mask as part of testing. The full figure is available upon request to the coresponding author or via the following link*. https://www.dropbox.com/sh/nkg2k1gu4aih3wz/AADy_o3CqlovLpCuXTE44uCYa?dl=0

#### Fluorescent Aerosol Dye Challenge

To assess the impact of improved mask fit on protection from external aerosols, the 3PM and UFM were challenged with fluorescent droplets (Fig. 4). Exposure to aerosol droplets without any protection resulted in coverage of 77 ± 7 % of the mannequin surface (Fig. 4B), and 3PM masks reduced the dye-covered area by 22%, (Fig. 4B) (P < 0.05). Dye accumulation was observed in the same zones of poor fit identified during thermal imaging. This observation demonstrates that these areas of poor fit provide an opportunity for dangerous aerosols to bypass the mask filter material. The UFM was three times more effective at reducing aerosol ingress compared to the 3PM (Fig. 4C) (P < 0.0001), even though the hard plastic of the mannequin reduced the ability of the UFM to form the tight seal normally generated as the mask is pulled against compliant skin by the adjustable tethering device.

**Figure 4.**
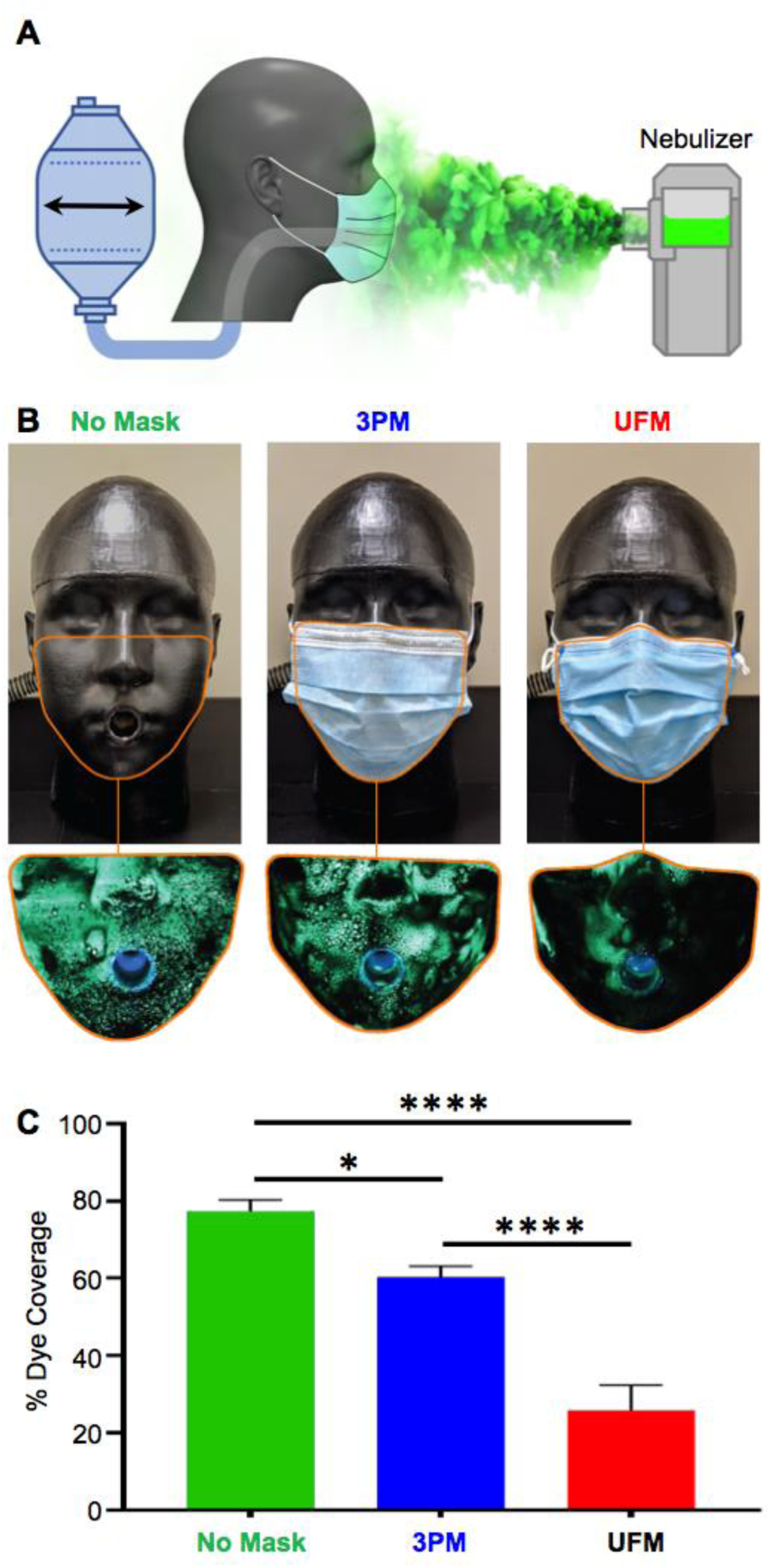
Aerosol Dye Examination of 3-ply Mask (3PM) and Ultra Fit Mask (UFM) (A) Schematic of the experimental setup where an aerosolized fluorescent dye was nebulized in the vicinity of the headform, with exhalation and inhalation breathing simulated by a modified resuscitator. (B) Fluorescent droplets deposited on a NIOSH medium headform with no mask, 3PM, and UFM. For scale, each image measures ca. 18 cm wide. (C) Relative exposure of the mannequin to aerosols assessed by the relative area covered by aerosolized fluorescent dye. Results represent the mean ± SD of 6 repeated trials. ^*^ and ^****^ indicate adjusted p<0.05 and p<0.0001 compared via a one-way ANOVA with Tukey post-hoc test.

### Quantitative Fit Test

We performed quantitative fit testing using a PortaCount® fit tester. 13 volunteers underwent a fit testing procedure per 29 CFR 1910.134, and 3PM, KF94 and UFM samples were evaluated (Fig. S2). The average UFM fit factor (FF) was 12.9 ± 7.2 (Fig 5A), which was 320% of the 3PM FF (FF = 4.0 ± 1.8, P < 0.001) and 240% of the KF94 mask FF (FF = 5.5 ± 5.9, P < 0.001) respectively, while the observed difference between the 3PM and KF94 mask FFs was not statistically significant (P > 0.9999) (Fig. 5A). During the ‘Bending Over’ exercise, all three masks had a lower FF, and the UFM FF did not show a significant improvement compared to the other two masks (Fig. 5D, P > 0.05). Bending-over involved moving the upper body while a hose was connected to the mask during the test. Facial leakage was introduced due to the large body motion and strain created by the experimental setup in this position. Three experienced UFM users, whom as test users didn’t require donning instructions, scored an average overall FF of 23.7 ± 5.5 (P < 0.001) whereas novice UFM users scored an average overall FF of 4.7 ± 2.5, suggesting further face fit improvement with experience (Fig. 5C). Interestingly, in novice users the UFM FF was 2.5 times higher than the 3PM FF (P < 0.005), consistent with a marked improvement in performance, even in novice users. While, in our user group, the presence of facial hair reduced fit factors, which was consistent with literature values (Fig. 5D), the UFM FF was still significantly higher compared to 3PM FF for users, both with and without facial hair (P < 0.001 and P < 0.0001, respectively). Regardless of familiarity, facial hair, or body movement, the UFM group significantly outperformed the 3PM and the KF94 groups.

**Figure 5.**
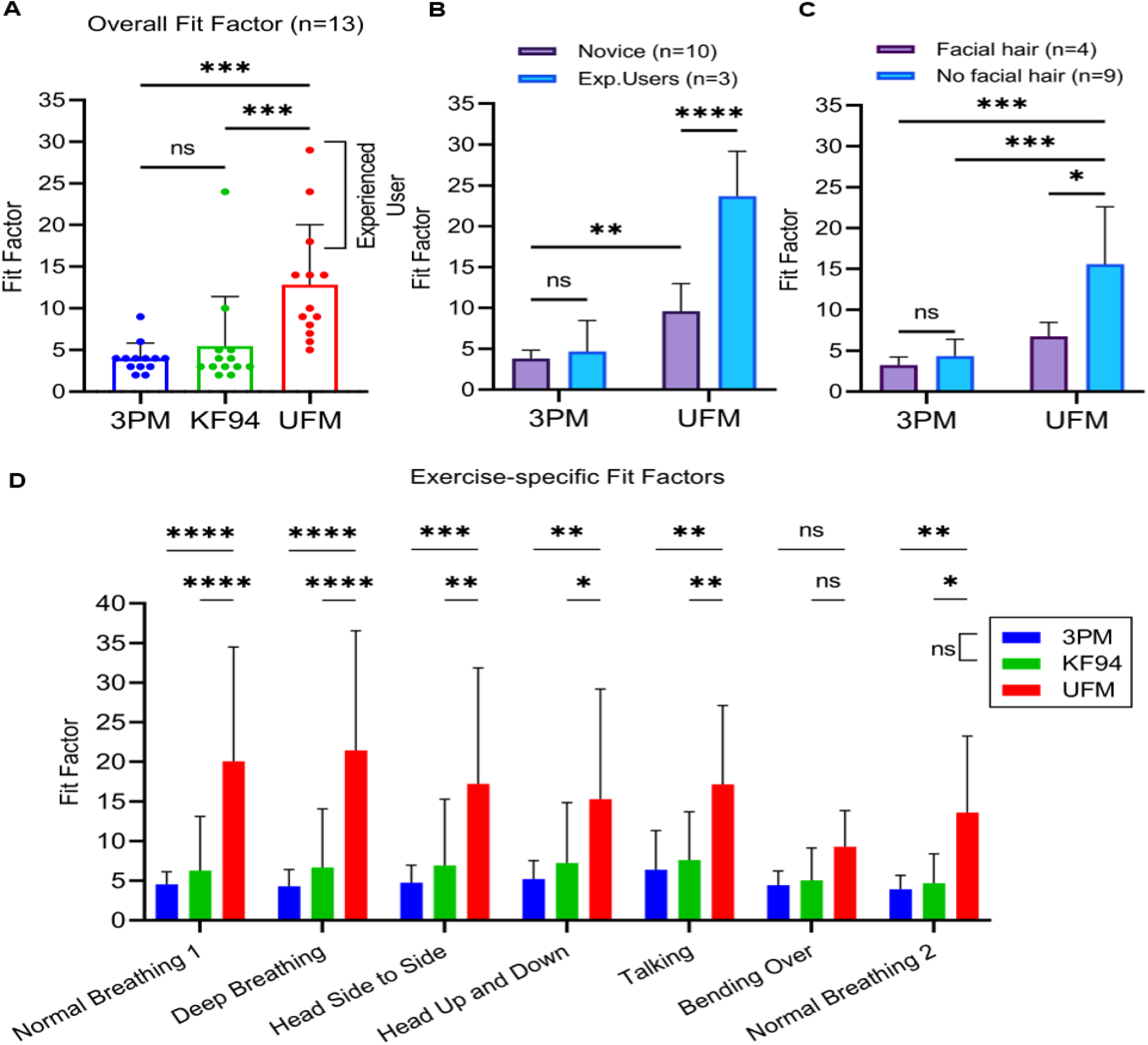
Quantitative Fit Test Results of 3-ply Mask (3PM), KF94 and Ultra Fit Mask (UFM) (A) Overall fit factors of quantitative fit test by Portacount 8038 for n=13 volunteers with 3PM, KF94 and UFM. (B) Fit factors between novice (n=10) and expert users (n=3). (C) Effects of facial hair with 3PM and UFM groups. UFM with no hair outperformed both 3PM with and without hair and as well as UFM with hair. was used. (D) Exercise-specific fit factors measured with Portacount 8038 of 3PM, KF94 and UFM. After reading a provided rainbow passage (‘Talking’), a non-measuring grimace exercise was performed per the OSHA fit test protocol. The post-grimace normal breathing exercise was labelled as ‘Normal Breathing 2’ to differentiate from the earlier normal breathing (‘Normal Breathing 1’). Bars are of the mean with error bars indicating the standard deviation. A Friedman test was performed for statistical analysis. ^*, **, ***^, and ^****^ indicate adjusted p-values of less than 0.05, 0.01, 0.001, and 0.0001, respectively.

### User Evaluation

To assess ease of donning and perceived comfort, we conducted a survey of individuals that had previously used the UFM. Each user had thus viewed the donning instructions and had experience using the UFM (Fig. 6A). Most responded that the UFM was easier or similar in difficulty to don compared to conventional 3PM, with only 13% reporting more difficulty donning the UFM (Fig. 6B). More than 90% of the respondents found it more comfortable than or similar to a 3PM disposable mask (Fig. 6C). With appropriate instruction provided, ease-of-use for UFM was comparable to the 3PM.

**Figure 6.**
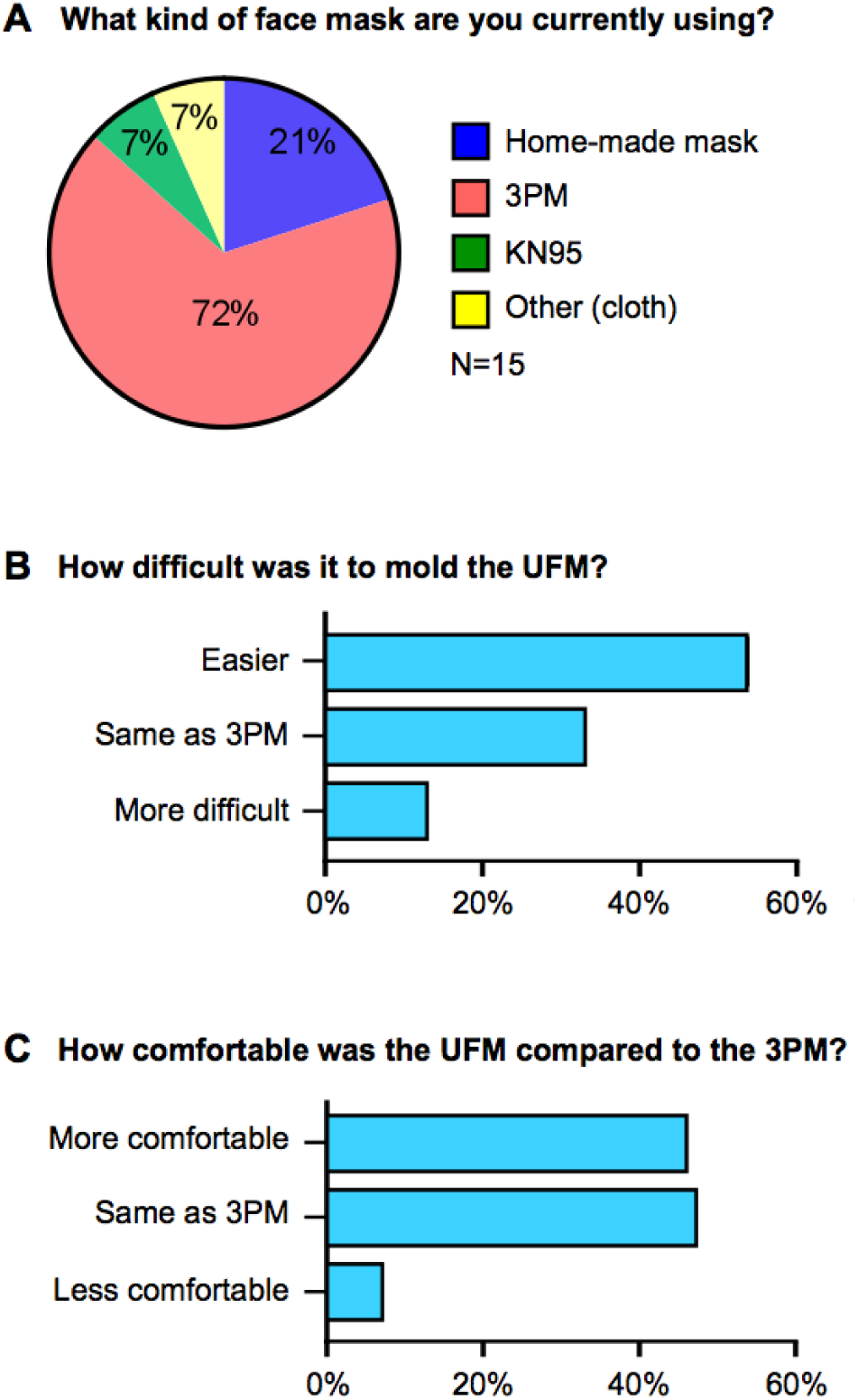
User Validation Survey with 15 Participants. (A) A majority of respondents (72%) used a conventional 3-ply mask (3PM). (B,C) >50% of respondents replied ‘equal or easier to don’ an Ultra Fit mask and ‘equal or more comfortable’ to wear an Ultra Fit mask compared to the 3PM.

### UFM Filtration efficiency and reusability

To evaluate if UFM masks filtration efficiency and ability to be reused after washing we evaluated filtration efficiency and air flow resistance. Filtration efficiency for the base UFM before washing was 88.2 ± 1.8 % with 59.8 ± 2.3 Pa of resistance to airflow (Fig. 7 A,B). As expected, machine washing diminished filtration efficiency by 40% and this corresponded to reduced resistance, indicating damage to the filter media during the wash (Fig. 7C). Hand washing at temperatures known to deactivate coronavirus (16) without detergents had no statistically significant effect on mask filtration or resistance to airflow. However, These results indicate that the UFM achieves a high filtration efficiency and is likely able to be sanitized and reused.

**Figure 7.**
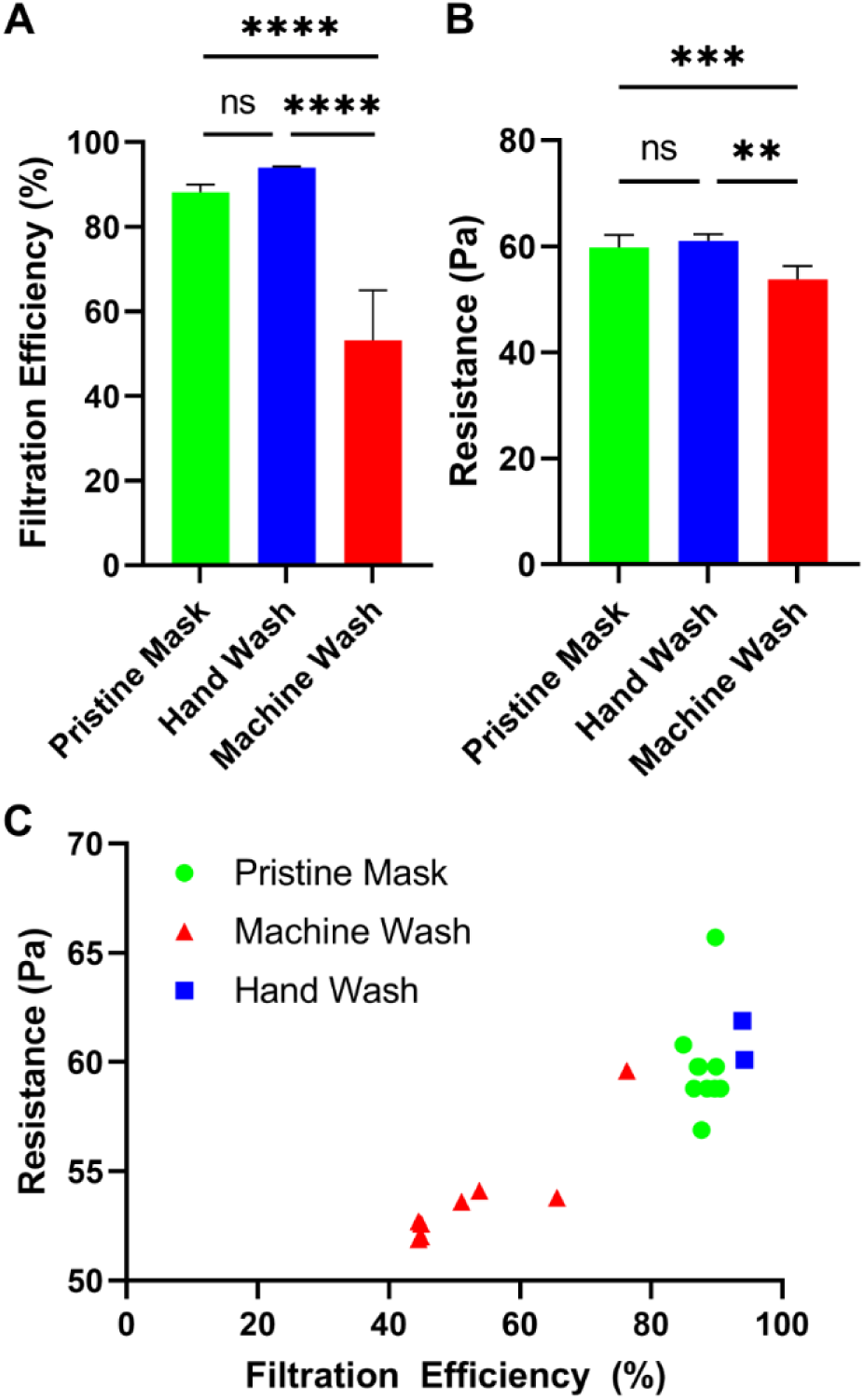
Filtration, Airflow, and Washability of the Ultra Fit Mask (UFM) (A) Filtration efficiency of the baseline (as-is, from packaging) UFM mask (n=10), and after hand-washing (n=2), and machine-washing (n=8). B) Resistance to airflow through the mask before and after the different washing methods. Plots represent the mean ± SD of unique test samples. C) Scatter plot showing the distribution and clustering of individual test samples based upon the filtration efficiency and resistance. ^**, ***, ****^, and ns indicate adjusted p<0.01, p<0.001, p<0.0001, and no significant difference (p>0.05) compared via a one-way ANOVA with Tukey post-hoc test.

### Modeling UFM impact on Community Spread on an airborne pathogen

To evaluate the potential impact of the UFM on the risk of infection in the context of a classroom and mass transit, we used a model of aerosol disease transmission using parameters obtained from the current COVID-19 pandemic. (17) Masking was beneficial in the model no matter the quality of the mask, but as fit and filtration efficiency increased the amount of protection increased (Fig. 8A). At with 50% of the class using masks the cloth mask, 3PM, excellent cloth mask, UFM, and N95 respirator reduced the relative risk of airborne infection by 18%, 24%, 44%, 62%, and 73% compared to no masks being used, respectively (Fig. 8A). We calculated the relative impact of each mask type on the airborne infection risk parameter, a predictive measure of the rate an individual is exposed to an infectious agent, for a subway car. We found similarly that the UFM out performed both model cloth masks and the 3PM (Fig. 8B). At 50% mask prevalence the UFM use dropped the airborne infection risk by 50.4% compared to the 3PM. In both model situations, the UFM approached the efficacy of N95 respirators to prevent the community spread of COVID-19.

**Figure 8.**
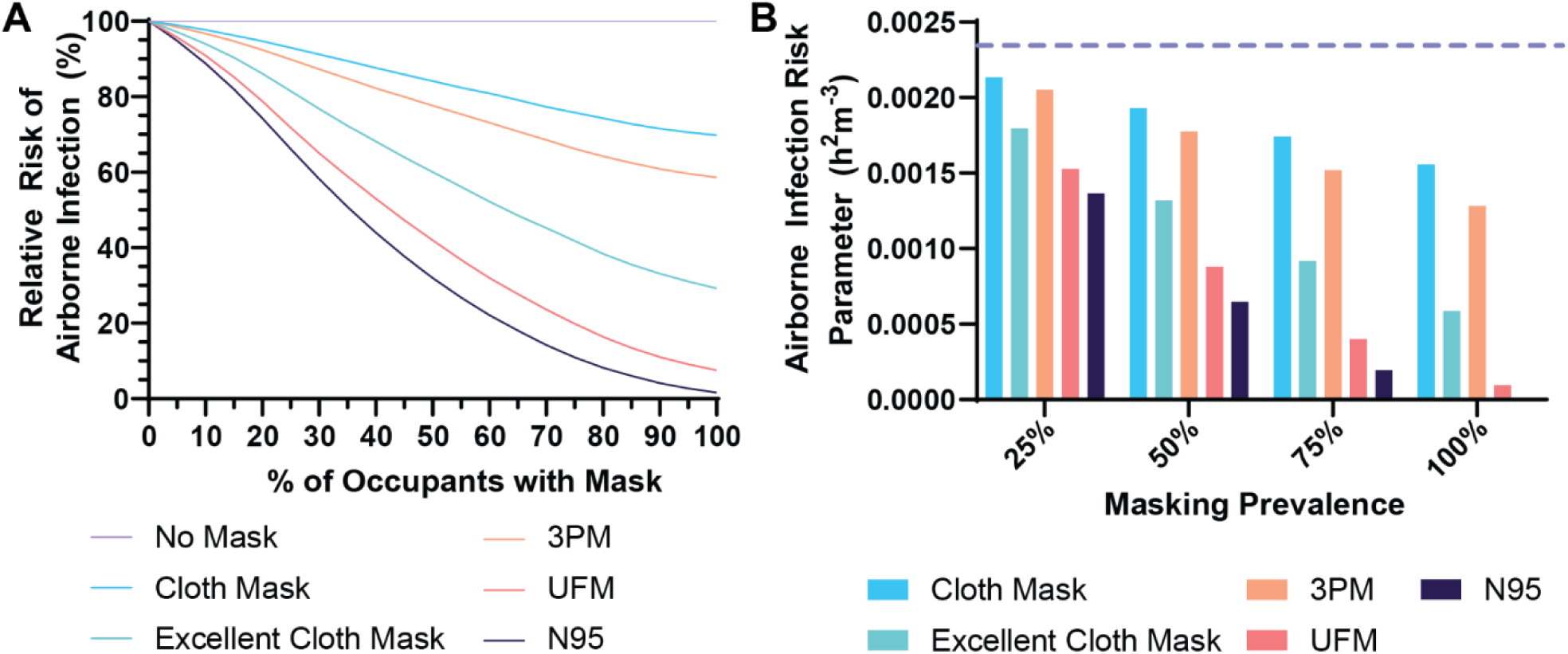
Model of Airborne Infection Risk by Mask Type and Usage. A) The relative risk of a new airborne infection occurring in a classroom type environment with varying rates of masking and mask types. B) The calculated individual infection risk parameter for individuals riding a subway train, assuming the presence of an infective individual. The dashed purple line indicates the risk parameter for no masking (17).

## Discussion

While improving face mask fit increases protection for both the community and individual (18), the most prevalent type of commercially available mask, disposable multi-layer face masks, such as 3PM, are plagued by poor fit issues (19). To address these persistent design challenges, we have developed a low-cost high-performance mask suitable for mass production and widespread community adoption.

From a series of comprehensive performance evaluations, including leakage visualization and measured FFs, we have shown that the UFM tripled the protection from aerosols compared to standard disposable face masks, for only $0.05 in additional material cost per mask. Each design element of the UFM was intentionally developed with a focus on manufacturability and scalability to enable the UFM to be produced on existing automated production lines with simple modifications to close peripheral gaps. As a result, large numbers of UFM can be produced at scale without the need for complex and expensive retooling. The UFM is a low-cost, mass-producible, easy-to-don, and comfortable face mask that minimizes face seal leakage by i) adding pliability to mask periphery to enable face molding to create a personalized facial fit, ii) integrating ear loop tension adjustment facilitating optimal compression of the mask on the wearer’s face, and iii) an inner “cough-trap” flap for blocking rapid respiratory jets induced by coughing or sneezing. With these three innovations, the performance of the UFM exceeds ASTM F3502 Level 2 Barrier Face Covering specifications and achieves the NIOSH criteria for a Workplace Performance Plus mask. While the quantitative fit experimental protocols implemented in the present study represent industry standards for evaluating face mask-fit performance (20), we performed additional experiments using thermal imaging, laser particle scattering, thermal imaging, and fluorescent photography visually to illustrate the ability of the UFM to prevent the expulsion and ingress of aerosols.

While much attention was paid to mask type during the COVID-19 pandemic, the experimental data suggest that absent proper fit, mask performance can be severely compromised. A study of the 9 most common respirator masks used by healthcare workers in France found that for five of the models, 92% of the units had FFs of less than 25 (21). In our study, all UFM users showed improved FF and experienced UFM users had an average overall FF of 23.7 ± 5.5, performance similar to that seen in widely used consumer grade respirator. In our study, UFM FF was superior to 3PM or KF94 masks, the performance of which was degraded by leakage, an observation consistent with previous studies (7, 22).

Throughout the COVID-19 pandemic, a wide range of alternative innovations designed to improve face mask fit have been developed. For example, face scanning technology was used to produce a user-customized mask brace, which is worn on top of the mask to minimize leakage (23). Despite the advantages offered through this approach, this innovative technology requires 3D printing of custom mask braces and is therefore challenging to deploy on a massive scale. Other notable innovations include the iMASC (24), a transparent sterilizable respirator, and a sew-free origami mask (25,26) that address face seal leakage issues by utilizing material’s natural properties. Elastic bands have also been developed to help seal masks but comfort and the need for additional aftermarket elements have limited their adoption (27). While all of these examples are valuable innovations that improve individual protection, it is presently uncertain if these designs can be produced at the scale necessary for widely deployed community respiratory protection which will require deployment of thousands of units.

Mask design requires optimization across multiple performance domains, including filtration efficacy, breathability, leakage, comfort, sustainability, cost, and supply chain robustness, with the added challenge of uniformly high performance across a wide range of face sizes and shapes. We assessed the capacity of our mask fit system (peripheral moldable elements, integrated elastics tensioners, and inner flange) to augment the performance of conventional 3PM, based on the knowledge that (1) conventional 3PM are widely available and (2) ASTM-F2100-compliant 3PM have been shown to have excellent filtration performance characteristics (28), implying that remediating face fit deficiencies in these masks would be most likely to produce a high-performing mask suitable for widespread community adoption. For the present study, quantitative fit testing was limited to the methods described in the OSHA fit test protocol for evaluating mask fit, and while these methods cover a wide range of scenarios that are reasonably representative of expected working activities, they do not include leisure activities such as jogging.

In addition to mask efficacy, mask-wearing compliance and reusability are important considerations for community masking initiatives. Reuse of disposable masks has been explored in the context of the mask shortages that emerged during the COVID-19 Pandemic. (29) As expected, we found that standard maschine washing deteriorated mask performance.(30) However, we were able to show that a simple hand wash method, using conditions that inactivate viruses, was able to preserve the filtration efficiency of the UFM. However, additional work is required to evaluate how users may be able to repeatedly sanitize the UFM for multiple use cycles. Compliance, as with all behavioral modifications, is a complex multifactorial challenge that extends beyond the properties of any individual mask. Additionally, the ability to reuse masks can dramatically reduce waste and environmental burden generated by mask mandates (31). While future work to validate the utility of our design elements in conjunction with reusable cloth face masks may be of benefit in specific community settings, it should be noted that cloth masks are well-known to offer lower, inconsistent filtration performance compared to meltblown non-woven filters in their 3PM alternatives (32).

The N95 respirators are poorly suited for community protection and are not user friendly. The tight fit of N95 masks creates discomfort and pressure injuries with extended and repeated use particularly during the COVID-19 pandemic (9). A survey of 4,306 clinicians in China during February 2020 found that 42.8% had experienced skin injuries due to their personal protective equipment (10). Respiratory protection provided by the N95 respirators come at the price of a compliant user-seal check and either qualitative or quantitative. Qualitative fit test based on human taste sensory is a realistic fit test program at healthcare providers. If no fit test was performed, the N95’s protection, due to inadequate face seal, is significantly reduced, and N95 respirators frequently need to be readjusted as 10-70% of initial tests fail to meet required levels depending upon the mask model and institution. (21) Also, access to N95 respirators was very limited to the public during the pandemic, due to the needs of healthcare personnel operating in high risk environments. These issues with N95 respirators, are part of the reason why even after shortages have somewhat resolved they still lack adoption in the broader community in low risk settings and would be difficult to rapidly deploy for use by the general population. The value comparison of the Ultra Fit mask, the N95 respirator and alternative devices (Table S1) shows that the UFM can be the best public respiratory protection. In models of community spread, the UFM approached the same level of protection as N95 masks to prevent the spread of airborne disease on a community basis. However, in an individual context, a fit-tested N95 still offers superior protection for use in aerosol-generating environments, and this protection exceeds the risks of discomfort and skin injury (9).

We used multiple methods to illustrate improved fit and respiratory protection with the UFM. While we developed the doable elements with the intent of achieving as close to a universal fit as possible, validation is still needed. We were limited in the variety of face shapes used in our assessments and future work is needed to demonstrate the degree to which UFM modifications enable improved fit for a variety of face shapes. Our modeling of individuals posed by the masks is limited by many of the assumptions specified describing our methods that were necessary to facilitate calculation. Additionally, modeling assumed that all COVID-19 infection occurs via aerosol transmission neglecting the risk of deposition on mucous membranes and contact transmission from the hands, which contribute to the spread of COVID-19 (33). In both the subway and classroom scenarios there was assumed to be exactly one contagious individual present, constant uniform spacing among individuals, uniform distribution of infectious particles within the space, and proper mask utilization.

However, these considerations do not accurately match reality, where there may not be no infecteous or potentially multiple infectious individuals present.

To the best of our knowledge, this work represents the first mass-producible barrier face covering achieving the NIOSH Workplace Performance Plus criteria enabled by three design elements in a highly-scalable face mask design. In combination with current face mask materials, the UFM design features significantly improved mask performance at low cost, reducing both the emission and ingress of particles and aerosols. In the context of community protection from the spread of airborne disease, the UFM approached the protection provided by the N95 respirator.

## Materials and Methods

### Ultra Fit Mask Fabrication

Materials used in the UFM are readily available and inexpensive. For the base mask, a 3PM with 3 pleats was selected (SupplyAID, SnowJoe LLC). This 3PM is compliant with GB/T32610-2016. However, any rectangularly-shaped pleated face mask and medical-use surgical mask can be used as a base mask. Segments of Zinc-galvanized steel wires with a diameter of 0.023” (8872K16, McMaster-Carr) were hot-melt glued on the four edges on the rear side of the base mask, which were then taped over with medical tapes for soft contact with the skin (Micropore™, 3M, MedFix, Medline). Empiric experimentation across a range of different wire diameters demonstrated the best performance with 0.50mm to 0.58mm bare wire. The material of the internal flap was the same material as the inner base mask layer, spunbond polypropylene nonwoven fabric with a weight of 30 – 40 grams per sq. meter. Custom-designed tethering adjusting components were cut from a stock polypropylene sheet (ITW Formex-GK17) and bonded with hot-melt glue on the upper corners on the front side of the base mask. The bulk cost of materials is estimated to be less than $0.05 per mask. At the time of writing, we have illustrated that these modifications can be performed with ultrasonic welding, eliminating the need for adhesives or tapes.

### Visualization of Face Seal Leakage

For visualization of exhaled vapor, a custom laser scattering setup was devised. An e-cigarette was used to produce vapor particles with a mean particle size ranging from 174 to 236 nm (34). Before donning a mask, the user inhaled vapor, donned the mask, and then exhaled. Particles were assessed using light scattered from a green laser pointer (Laser 303, λ_c_ = 532nm, P_max_ <5mW) modified with a cylindrical lens (LK1395L1, Thorlabs, Inc.). The exhalation event was recorded on an Apple iPhone × mounted on a tripod. For relative scattering analysis, the green channel of video files was analyzed over time using MATLAB code. Relative scattering was calculated as a sum of pixel values divided by the segmented area. Plots were then normalized to the theoretical maximum.

For infrared thermal imaging, two cameras, T440 (FLIR Systems) and CompactPro (Seek Thermal) were initially used. For convenience of data transfer, CompactPro was used for image acquisition. The subject was seated on a chair, and during imaging, the room was air-conditioned at approximately 20°C. For plotting thermal variation over time, 5-by-5 pixel regions were segmented for the left and the right and a 20-by-20 pixel region was segmented for the center. Mean values of the segmented regions were plotted over time. Thermal video data was acquired at 8.6 frames/second.

A single-mirror Schlieren optics setup to visualize refractive index changes in the air caused by changes in density, temperature, and pressure was set up by the Harvard Natural Sciences Lecture Demonstration Team. Due to inactivity in the building, however, the lecture hall was not air-conditioned for the experiment, so the obtained image contrast was lower than what one might normally expect.

An aerosol dye challenge was used to evaluate the protection provided by the mask to external aerosol exposure. The aerosol dye was generated 20 cm away from a head model, which was 3D printed using data obtained the National Institute of Occupational Health and Safety (NIOSH) Anthropometric data and ISO headform (35), and was outfitted with a 2 cm tube connected to the mouth. To simulate inhalation and exhalation a 2000 ml resuscitator bag (Vodeson Inc.) was modified by closing the one-way valves on the back of the bag, and removing the valve at the interface with the mouthpiece. The resuscitator bag was physically compressed and then allowed to expand in 3 second cycles over a 30 second test period to simulate breathing. Aerosol particles (27.94 ± 18.96 µm in diameter, 483,000 ± 102,000 particles per second, mean ±SD) of fluorescent dye (EC6 RECOLOUR Dye UV Green, XSPC) were generated from a model MN-01A ultrasonic mesh nebulizer. Dye on the surface of the mannequin was imaged using a 365 nm ultraviolet light source (Model UVGL-25) and a digital camera. The percent of the mannequin head’s surface coated in dye underneath the face mask was determined using image analysis in ImageJ 1.53E (National Institute of Health).

### Quantitative Fit Test

For a quantitative fit test, a commercial PortaCount® Respirator Fit Tester 8038 was rented (Raeco Rents, LLC) for the duration of experimentation. Automatic daily checks were performed before use. A standard protocol per 29 CFR 1910.134, which consisted of 8 exercises, was employed. After five exercises comprising different breathing styles (normal and heavy), head motions (side-to-side and up-and-down), and reading a provided passage, a wearer was asked to grimace to intentionally break a face seal, followed by bending over towards the toes before the final normal breathing was attempted. A fit factor was automatically computed and displayed at the end of each exercise except for non-breathing ‘grimace’ exercise. An overall fit factor was automatically computed at the end of the whole exercise cycle. With N95 Companion mode enabled, PortaCount® 8038 discriminates the contribution of leakage through the filter by use of electrostatic classifier and counts only particle sizes in the range of 25 nm to 60 nm. Therefore, the fit factors, as a measure of face seal leakage, are the ratio of ambient particle concentration outside the mask to that inside the mask for particle sizes between 25 nm and 60 nm. A conventional 3PM disposable mask and KF94 (AirQueen Nano, Soomlabs), a Korean standard mask rated to filter 94% of particles larger than 0.3 microns in size (36), were used as reference controls (Fig. S2). The 3PM used in our experiment was sold as SupplyAID by SnowJoe LLC, which also served as the base mask for UFM.

### User Validation

An online survey form was created and circulated for collecting user feedback. Questions asked in the survey were multiple-choice questions and short questions about what type of face mask a user wears, and the user’s assessment of the Ultra Fit mask. Full questionnaires are available at https://forms.gle/h4UnPkkn8aeCQQdT6

### Mask Washing Methods

Masks were tested at SGS North America Inc. All base mask materials were from the same lot (New Normal Surgical Mask, Medifac, Korea). The hand-washed and machine-washed samples were tested at UMass Lowell & Advanced Functional Fabrics of America. At both facilities, the same automated filter test apparatus, TSI 8130 (TSI Inc.), was used per modified NIOSH TEB-APR-STP-0059 at a flow rate of 60 lpm as flat media. Conditions for washing were selected based upon processes that had shown promise through heat inactivation of coronavirus and preserving mask function (16,37,38). For hand washing, the sample was soaked in 65° C of plain water without detergent for 10 minutes and dried with a home-appliance dryer at 76° C for 20 minutes. For machine washing, a home-appliance washing machine was used with hot plain water without detergent and cold rinsing. To dry the mask was spun for for 11 minutes followed by an auto-timed 3-minutes drying at 35° C.

### Modeling Airborne Infection Risk Mitigation

To evaluate how the design modification to the UFM would influence the risk of airborne pathogen spread we used the computational model developed by Peng. Et. al. (17) Briefly, the model uses the concept of an enclosed box to estimate the amount of aerosol contamination taking into consideration: ventilation, volume of the space, and the amount of virus put into the air by infected individuals (factoring in mask leakage). The model then calculates an airborne risk parameter that takes into account the amount of virus in the air over the exposure period, the volume of air uninfected individuals take in during breathing, and mask efficiency. The Wells-Riley model of infection is then used to calculate the probability of an individual becoming infected after exposure.(39) We derived the values for the mask efficiency for the UFM based upon the results of the quantitative fit testing, filter testing, and the aerosol exposure test. For the cloth mask and excellent model cloth, we used the lowest and highest values for the reported ranges, respectively, for Eikenberry, et. al.(40) The mask efficiency values entered into the calculation tool generated in Peng. et. al. and the fraction of people with masks was varied systematically. The baseline settings for ventilation, volume, duration of stay, activity adjusted breathing rates, and infectious quanta for COVID-19 within the classroom and subway environments, were used. The number of individuals in the classroom and subway car were set to 20 and 35, respectively, and there was assumed to be one infective individual in each case. Data in the classroom was normalized against the risk of a secondary infection occurring in the absence of any masks over a 50 minute class period. As the duration of ride on a subway is highly variable, we calculated the absolute infection risk parameter, which is independent of the duration of the ride, at 0, 25, 50, 75, and 100% mask utilization.

## Statistical Analysis

Data processing and statistical analysis were performed using a combination of MATLAB, ImageJ, and GraphPad Prism 9.0. All graphs and reported values represent mean ± standard deviation unless otherwise stated. A Shapiro-Wilks test was used to test if data were normally distributed before selecting parametric or nonparametric tests. Comparisons across multiple groups were performed on nonparametric data using Kruskal-Wallis for multiple groups and Mann-Whitney for paired tests. For parametric data, a one-way ANOVA was used. A Tukey post-hoc test was applied to correct for multiple comparisons and an adjusted p-value of < 0.05 was used as the threshold for significance. The specific test used is described in the caption of each figure.

## Data Availability

The data described in this manuscript are available through request to the corresponding authors.

## Acknowledgements

We acknowledge funding from Katharos Laboratories, LLC. We’d also like to thank all of the volunteers, Harvard University undergraduate students, the Harvard Natural Sciences Lecture Demonstration team, Harry Miller Co., early users for their design feedback (Alice, David, Nick, and Laura), and the Mass General Brigham Center for Covid Innovation, Susan Goldsmith, and Max Miller for their contributions. This work was also supported by the Incheon National University Research Grant in 2021 to KY.

## Author contributions

Conceptualization: CH, SJG, AS, JMK

Methodology: CH, JCW, MMJ

Investigation: CH, KS, MMJ

Visualization: CH, JCW, MMJ

Supervision: AS, JMK

Writing—original draft: CH, KS, XW, MMJ

Writing—review & editing: CH, KS, XW, YK, JCW, AS, MMJ, JMK

## Competing Interests

The UFM technology described herein has been optioned to Katharos Labs, a company in which Jeffrey M. Karp, Anthony Samir, Steven J. Gordon, and Chulho Hyun hold equity. J.M.K. has been a paid consultant and or equity holder for multiple companies including Katharos Labs (listed here https://www.karplab.net/team/jeff-karp). The interests of J.M.K. were reviewed and are subject to a management plan overseen by his institution in accordance with its conflict of interest policies.

## Data and materials availability

The data described in this manuscript are available through request to the corresponding authors.

## Notes

### Funding Statement

We acknowledge funding from Katharos Laboratories LLC. We would also like to thank all of the volunteers, Harvard University undergraduate students, the Harvard Natural Sciences Lecture Demonstration team, Harry Miller Co., early users for their design feedback (Alice, David, Nick, and Laura), and the Mass General Brigham Center for Covid Innovation, Susan Goldsmith, and Max Miller for their contributions. This work was also supported by the Incheon National University Research Grant in 2021 to KY.

### Author Declarations

Mass General Brigham IRB

