## Supplemental Information for "The Ultra Fit Community Mask - Toward Maximal Respiratory Protection via Personalized Face Fit"

### Supplementary Materials

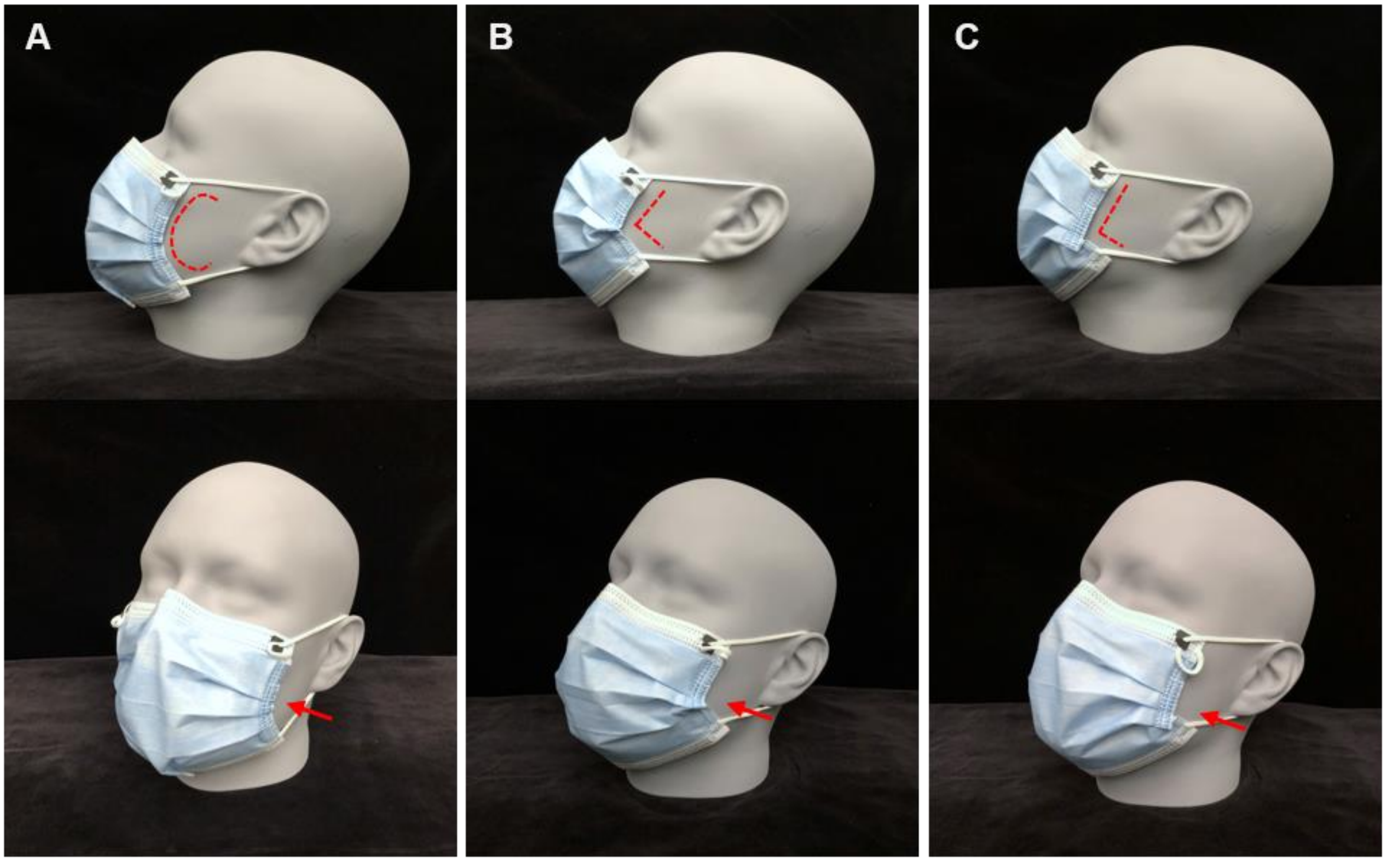


**Figure S1. Different Donning Styles with the Ultra Fit Mask for Improved Fitting.**

(A) C-shaped: C-shaped curve on the side edges with pinching the chin edge. (B) V-shaped: pinching the side wires resembling a V. Optional chin pinching may further tighten fitting depending on the user's preferences. (C) L-shaped: folding the lower parts of the side wires resembling an L. Optional chin pinching may further tighten fitting depending on user's preferences.

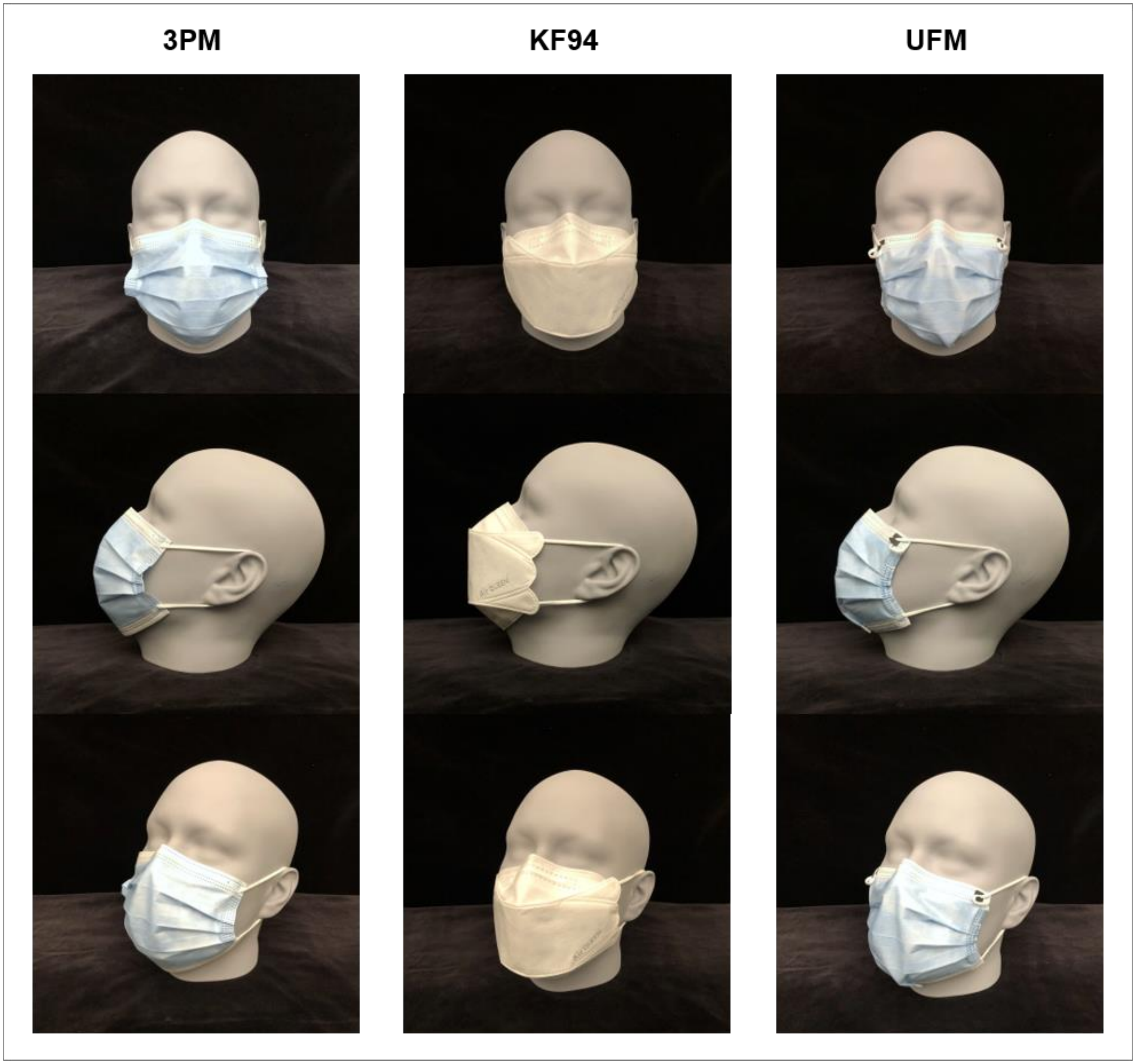


**Figure S2. Three Different Face Masks (3PM, KF94, UFM) Used for Quantitative Fit Test Donned on a ISO 16976-2 Medium Headform.**

(A) 3PM. Large gaps on the cheek sides are clearly visible. (B) KF94. Side gaps are visible. Infraorbital-nasal region and the chin appear closed. (C) UFM. The mask showed reduced side gaps and was tucked under the chin by pinched chin wire.

Figure S1.mp4

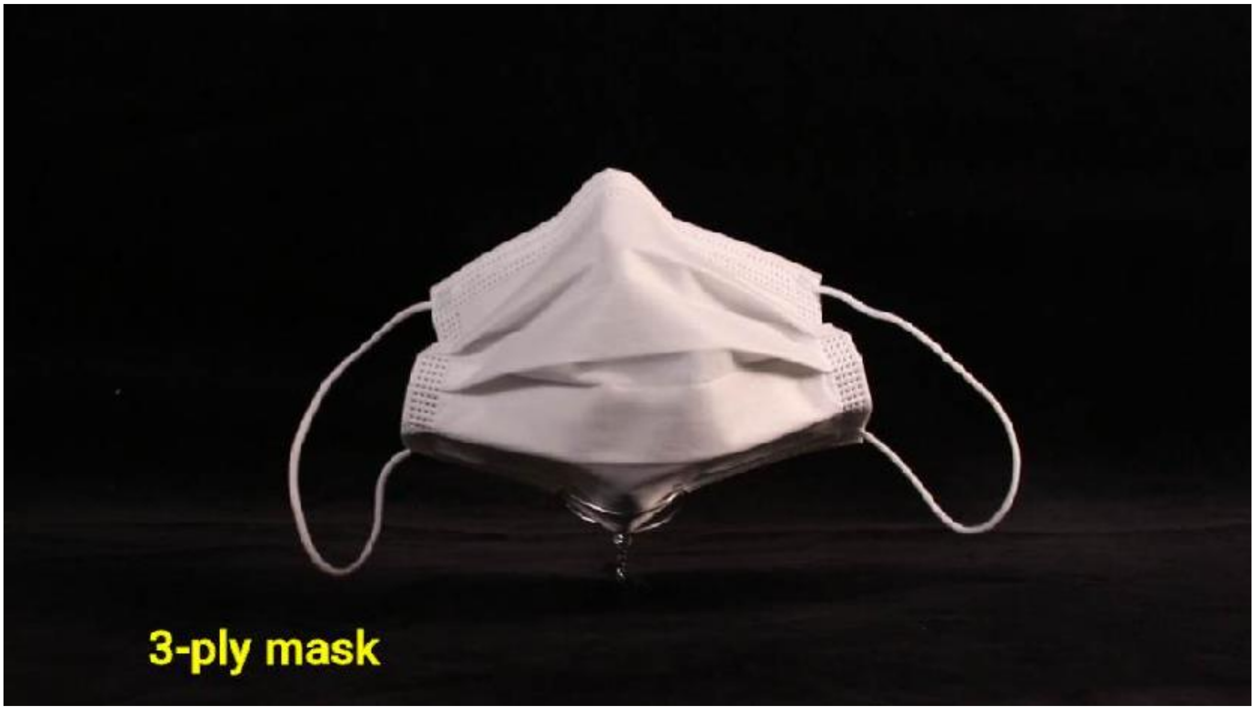


**Movie S1. Movie of 3-ply Mask and Ultra Fit Mask.**

Masks on a rotating turntable. Differences in structural rigidity are clearly visible.

Figure S3.mp4

**Movie S2. Movie of Exhaled Vapor Tracking of the Front and Side views.**

***NOTE:*** *Per medRxiv policy that manuscripts not contain 'photographs/videos and any other identifying information of people' we have removed movie S2 to comply with this policy. In the this items, an author of this manuscript can be seen wearing the mask as part of testing. The full figure is available upon request to the coresponding author or via the following link.*

https://www.dropbox.com/sh/nkg2k1gu4aih3wz/AADy_o3CqlovLpCuXTE44uCYa?dl=0

Figure S4.mp4

**Movie S3. Infrared thermal imaging movie with 3-ply mask and Ultra Fit mask.**

***NOTE:*** *Per medRxiv policy that manuscripts not contain 'photographs/videos and any other identifying information of people' we have removed movie S2 to comply with this policy. In the this items, an author of this manuscript can be seen wearing the mask as part of testing. The full figure is available upon request to the coresponding author or via the following link.*

https://www.dropbox.com/sh/nkg2k1gu4aih3wz/AADy_o3CqlovLpCuXTE44uCYa?dl=0

**Table S1: Value Comparison**

| | **Ultra Fit Mask** | **Conventional Mask**<br>(e.g cloth masks, 3-ply masks, KF94) | **Clinical Grade Respirators**<br>(e.g N95, KN95, FFP3) | **Alternative Solutions**<br>(e.g mask fitter, double-masking, knotting) |
|---|---|---|---|---|
| **Total Inward Leakage (excluding filter penetration leakage)** | ~8% | 20-40% (1,2) | < 1% (if fit test passed)<br>If fit test failed, 4% (3) | Can achieve < 1% in conjunction with disposable mask (4) |
| **Filtration**[a] | 88% (at 10cm/s), > 95% (at 85 lpm) NIOSH test method | 25-80% (5), specified at 95%[b] | > 95% (at 85 lpm) NIOSH test method | N/A |
| **Fit (Fit Factor, FF)** | 12.9 (n=13) | 2.6 - 4.4 (1,2) | >100 (if passed a fit test)<br>< 25 (if failed a fit test) | Help achieving mask FF >100 (4) |
| **Breathing Resistance** | 6.1 $mmH_2O$ (at 10cm/s)<br>5.1 $mmH_2O$ (at 85 lpm) | varies (4-27 $mmH_2O$)[c] | <35 mm H2O column (at 85 lpm) | High/Medium |
| **Comfort** | Good | Good | Poor | Medium or Poor |
| **Accessibility to public** | Good | Good | Poor | Good |
| **Manufacturing at scale** | Good | Good/Poor | Intermediate | Good or N/A |
| **Cost** | Low (~$0.50, $0.05 addition to base mask price) | High for cloth mask (>$0.50)<br>Medium for KF94 (~$0.50)<br>Low for 3-ply mask (<$0.50) | High (>$1.00) | High (>$1.00) |
| **Easiness-to-use** | Good | Good | Intermediate | Intermediate |
| **Washability/Resuability** | Yes | Cloth: Yes<br>Non-cloth: No | Yes (6) | Depends |

| | | | | |
|---|---|---|---|---|
| **Contact Dermatitis Risk** | No | No | Yes (7,8) | No |
| **Applicable Standards/Certification** | ASTM F3502 Level 2 for both filtration & breathability. NIOSH Workplace Performance Plus mask | Foreign standards e.g GB/T 32610-2016, YY/T 0969-2013, GB2626-2006, KF94, AFNOR SPEC S76-001 | Surgical N95 designation | N/A |

[a]NIOSH TEB-APR-STP-0059, “Determination of Particulate Filter Efficiency Level for N95 Series Filters Against Solid Particulates for Non-Powered, Air-Purifying Respirators Standard Test Procedure (STP)”

[b]Bacterial filtration efficiency per ASTM F2101 or equivalent foreign standards

[c]observed by the authors using TSI 8130 and NIOSH TEB-APR-STP-0059.

4. Fix The Mask Data —the Essential Mask Brace | by Fix The Mask — the technical. | Medium, (available at https://fixthemask.medium.com/fix-the-mask-data-the-essential-mask-brace-ad5bdf33f506).

5. P. W. Clapp, E. E. Sickbert-Bennett, J. M. Samet, J. Berntsen, K. L. Zeman, D. J. Anderson, D. J. Weber, W. D. Bennett, Evaluation of cloth masks and modified procedure masks as personal protective equipment for the public during the COVID-19 pandemic.

6. N95DECON, N95 Decontamination & Reuse Method Decision Matrix (2020).

7. J. Yu, J. K. Chen, C. M. Mowad, M. Reeder, S. Hylwa, S. Chisolm, C. A. Dunnick, A. M. Goldminz, S. E. Jacob, P. A. Wu, J. Zippin, A. R. Atwater, Occupational dermatitis to facial personal protective equipment in health care workers: A systematic review. J. Am. Acad. Dermatol. 84, 486–494 (2021).

8. A. Hornbeck, J. K. Cichowicz, S. Kilinc-Balci, D. Rottach, J. Pollard, H. Boyles, Skin Irritation from Prolonged Use of Tight-Fitting Respirators | Blogs | CDC (2020), (available at https://blogs.cdc.gov/niosh-science-blog/2020/08/04/skin-irritation-respirators/).
